# Ultrasound versus fluoroscopy as imaging guidance for percutaneous nephrolithotomy: A systematic review and meta-analysis

**DOI:** 10.1101/2022.10.13.22281046

**Authors:** Razman Arabzadeh Bahri, Saba maleki, Arman Shafiee, Parnian Shobeiri

## Abstract

**Objectives:** To determine whether the outcomes of ultrasound-guided percutaneous nephrolithotomy (UG-PCNL), an alternative to traditional fluoroscopy-guided percutaneous nephrolithotomy (FG-PCNL), are comparable.

**Methods:** A systematic search of PubMed, Embase, and the Cochrane Library was carried out to discover investigations comparing UG-PCNL to FG-PCNL, and accordingly, a meta-analysis of those studies was performed. The primary outcomes included the stone-free rate (SFR), overall complications based on Clavien-Dindo classification, duration of surgery, duration of patients’ hospitalization, and hemoglobin (Hb) drop during the surgery. All statistical analyses and visualizations were implemented utilizing R software.

**Results:** Nineteen studies, including eight randomized clinical trials (RCTs) and eleven observational cohorts, comprising 3016 patients (1521 UG-PCNL patients) and comparing UG-PCNL with FG-PCNL met the inclusion criteria of the current study. Considering SFR, overall complications, duration of surgery, duration of hospitalization, and Hb drop, our meta-analysis revealed no statistically significant difference between UG-PCNL and FG-PCNL patients, with p-values of 0.29, 0.47, 0.98, 0.28, and 0.42, respectively. Significant differences were discovered between UG-PCNL and FG-PCNL patients in terms of the length of time they were exposed to radiation (p-value< 0.0001). Moreover, FG-PCNL had shorter access time than UG-PCNL (p-value= 0.04).

**Conclusion:** UG-PCNL provides the advantage of requiring less radiation exposure while being just as efficient as FG-PCNL; thus, this study suggests prioritizing the use of UG-PCNL.

## Introduction

Urolithiasis is a common urological disease with increasing prevalence (1). Percutaneous nephrolithotomy (PCNL) is a minimally invasive procedure for removing complex or large kidney stones (2, 3) and is conventionally performed under fluoroscopic guidance. A major concern related to fluoroscopy-guided percutaneous nephrolithotomy (FG-PCNL) is the effect of exposure to ionizing radiation by patients, surgeons, and operating room personnel (4, 5). Thus, an alternative imaging technique for PCNL would be advantageous (6). To avoid radiation exposure, some surgeons prefer ultrasound-guided percutaneous nephrolithotomy (UG-PCNL) (7). Given the higher rate of availability of ultrasound (US) devices in most peripheral hospitals, the use of UG-PCNL also increases the number of PCNL procedures. Also, the total cost of UG-PCNL is 30% less than that of FG-PCNL in every case (8). PCNL is performed in prone, supine, or flank positions. The prone position is the preferred modality for creating percutaneous access and localizing stones during FG-PCNL (9) although, in obese patients, it is not an ideal position(10). UG-PCNL can also be performed in the flank or supine position with a lower risk of complications during anesthesia(11). There have been various studies, including observational studies or clinical trials, comparing the feasibility, safety, and efficacy of ultrasound-guided and fluoroscopy-guided PCNL, but the conclusion has scarce available data and the choice between these modalities is based on the preference of the urologist. Therefore, updating this data is mandatory. In addition, there has not been a comprehensive assessment of randomized clinical trials (RCTs) and observational cohort studies comparing UG-PCNL and FG-PCNL. Therefore, we aimed to systematically review and conduct a meta-analysis to compare the efficacy of UG-PCNL with FG-PCNL in different outcomes for the treatment of urolithiasis.

## Methods and materials

This study was conducted based on Preferred Reporting Items for Systematic Reviews and Meta-Analyses (PRISMA) guidelines. The protocol of this study is registered in PROSPERO (CRD42022327222)

### Search strategy

A comprehensive search was conducted in international databases, including Cochrane library, PubMed, and Embase, for relevant studies published from the inception to March 19th, 2022. The search was conducted again for the determination of newly published and relevant studies one week before the submission of the manuscript. The search keywords were categorized into three groups: Ultrasound, fluoroscopy, and nephrolithotomy. In the ultrasound group, we used any possible keywords such as ultrasound, US, ultrasound-guided, ultrasonography, and ultrasonographic. In the fluoroscopy group, we used all possible keywords, including fluoroscopy, X-ray, and fluoroscopic. In the nephrolithotomy group, the keywords used in the search strategy were percutaneous nephrolithotomy, minimally invasive percutaneous nephrolithotomy, and PCNL. The keywords were combined with “AND” between the groups, and with “OR” in each group.

### Eligibility criteria

The inclusion criteria for study selection were as follows: (a) patients with urolithiasis condition; (b) comparison of ultrasound-guided PCNL and fluoroscopy-guided PCNL; (c) reporting of at least SFR and complication rate, and (d) studies in the English language. The exclusion criteria for study selection were as follows: (a) non-randomized studies, (b) Meta-analysis studies, and (c) review studies. No limitation was imposed in this study for study sample sizes and patient characteristics.

### Data extraction and quality assessment

The initial screening of studies was carried out by two reviewers independently based on titles and abstracts to exclude non-related studies. The full text of related studies was then reviewed for confirmation of eligibility criteria meeting and data extraction. The data extraction of each study using an Excel-based sheet were checked and discussed by two reviewers independently. The data sheet included the first author names, type of studies, year of publications, number of patients in ultrasound and fluoroscopy group, patients’ characteristics, SFRs, PCNL techniques, multiple stone status, stone burden, hydronephrosis degree, ultrasound probe, sheath size, dilator, complication rate, surgery time and Hb decrease after the surgery. The methodological quality of the included studies was independently assessed by two reviewers using the national institute of health (NIH) quality assessment tool for cohort studies and the risk of bias (RoB2) method of the Cochrane Collaboration for RCTs.

### Outcomes

Five primary outcomes and five secondary outcomes were evaluated and analyzed. The primary outcomes included the SFRs, overall complications based on Clavien-Dindo classification, duration of surgery, duration of patients’ hospitalization, and hemoglobin (Hb) drop during the surgery. The secondary outcomes included need for blood transfusion, fever after the surgery, radiation exposure of the patients, time to access the stone, and the number of attempts by the surgeon for the procedure.

### Statistical analysis

The risk ratio (RR) was used to summarize the pooled effect size of dichotomous outcomes, and the standardized mean difference (SMD) was used for reporting the results of continuous outcomes. Study heterogeneity was assessed using the Chi-square test and I2 statistic, with I2 values of <25% indicating a low amount of heterogeneity. A fixed effect meta-analysis was performed in the case of low heterogeneity; otherwise, a random effect model was used. In order to investigate any potential effects of the type of study, sub-group analysis was carried out based on their design (whether a randomized controlled trial (RCT) or an observational cohort). Publication bias was assessed using funnel plots and Egger’s regression test for funnel plot asymmetry. We did not report publication bias for outcomes with less than ten studies included. A meta-regression was performed to investigate the possible association between the publication year as a measure of clinicians’ experience in performing US-guided PCNL and the effect size. Meta-regression analyses were only done on primary outcomes. All statistical analyses and graphics were carried out using R (version 4.1.3) (R Core Team, 2020) and the meta package.

## Results

### Study Characteristics

Nineteen studies, including eight RCTs and eleven observational cohorts comparing UG-PCNL with FG-PCNL, were included in our study (Fig1). The characteristics of the included studies are presented in Table1. The first study was published in 2008 and the majority of the studies have been published in recent years (2016-2021). The sample size of the articles ranged from 45 to 906. A total of 3016 patients were evaluated in the included studies (of which 1521 patients underwent UG-PCNL)

**Fig. 1:**
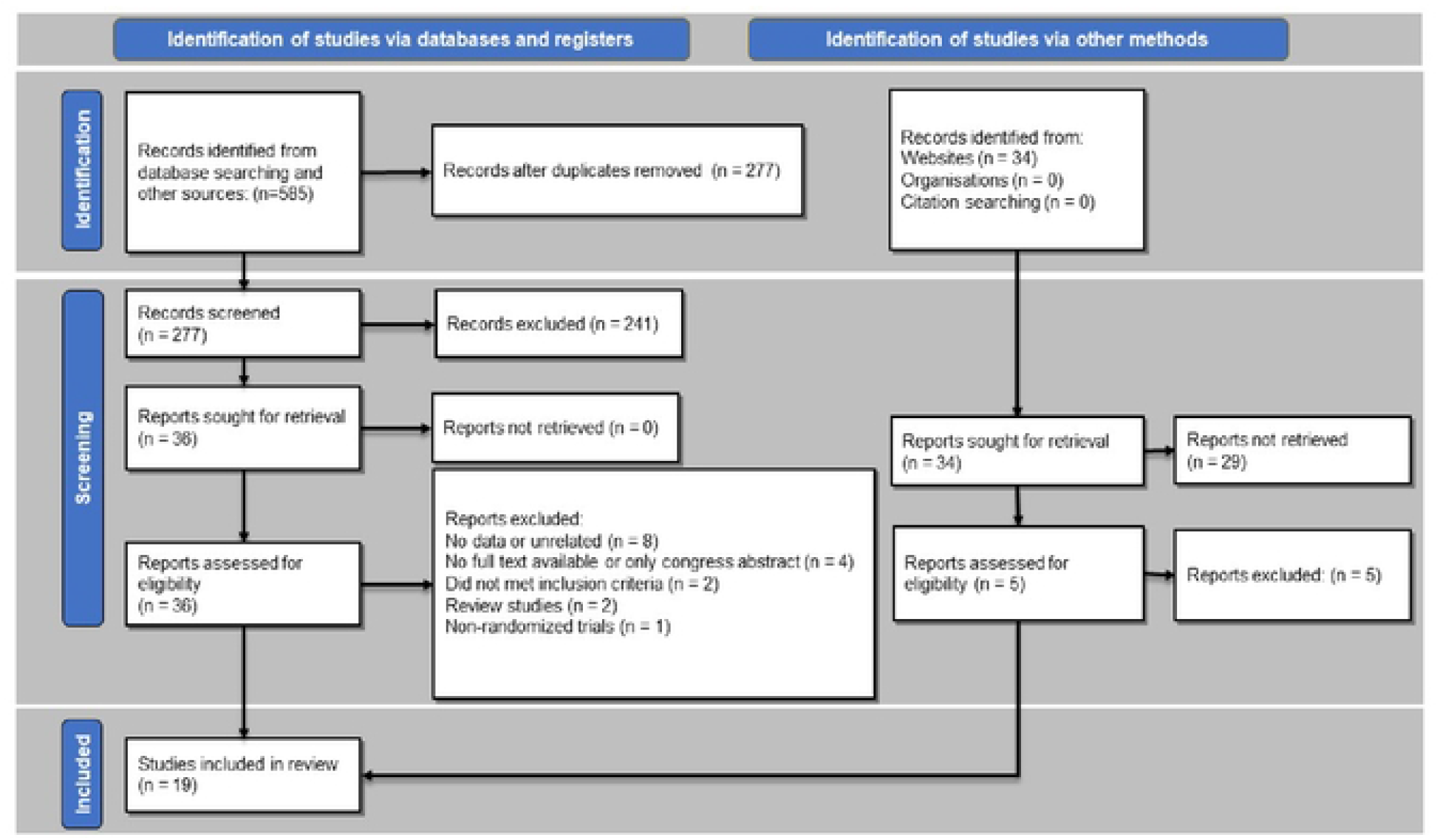
PRISMA flowchart of the literature search and selection of the articles.

**Table 1.**
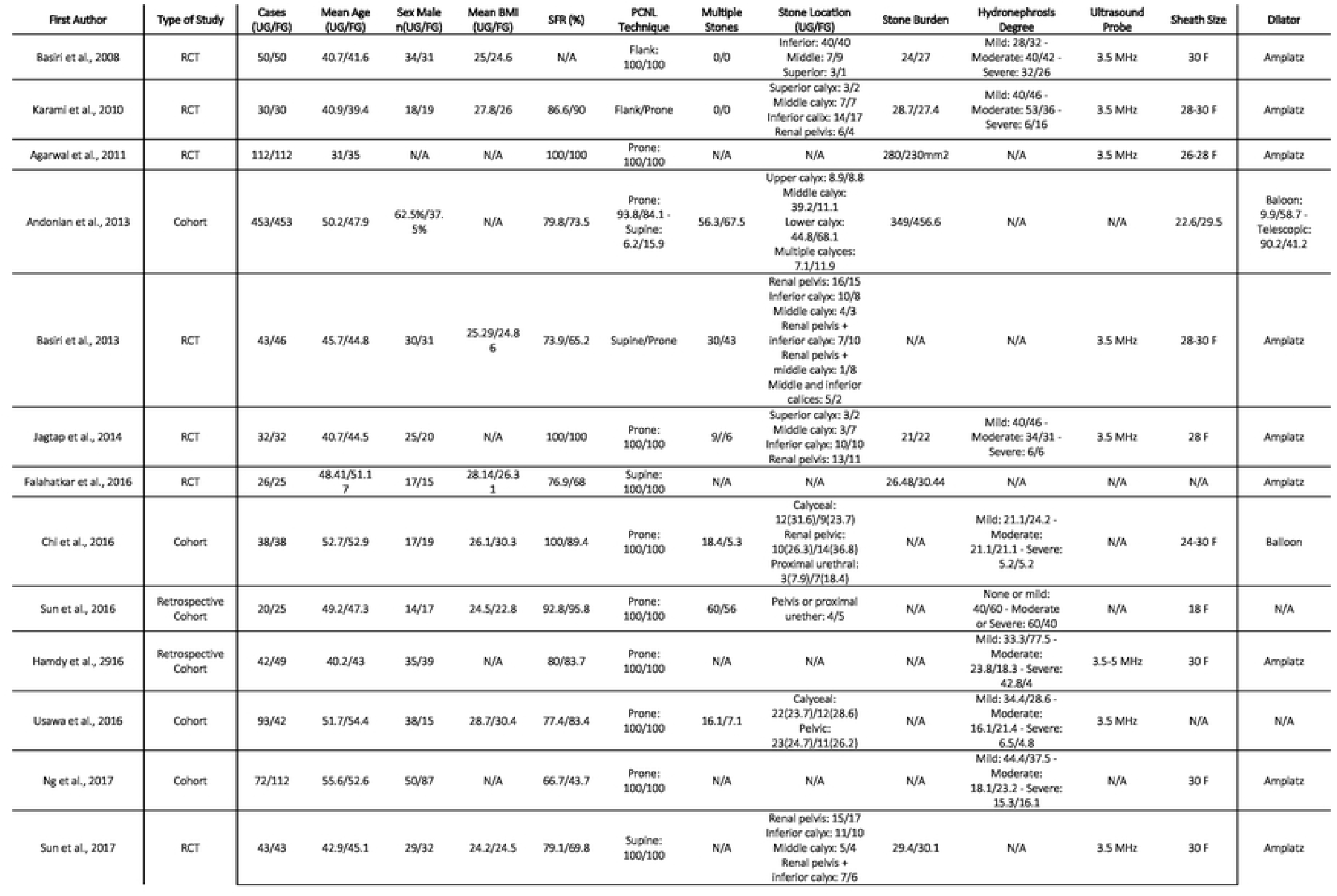
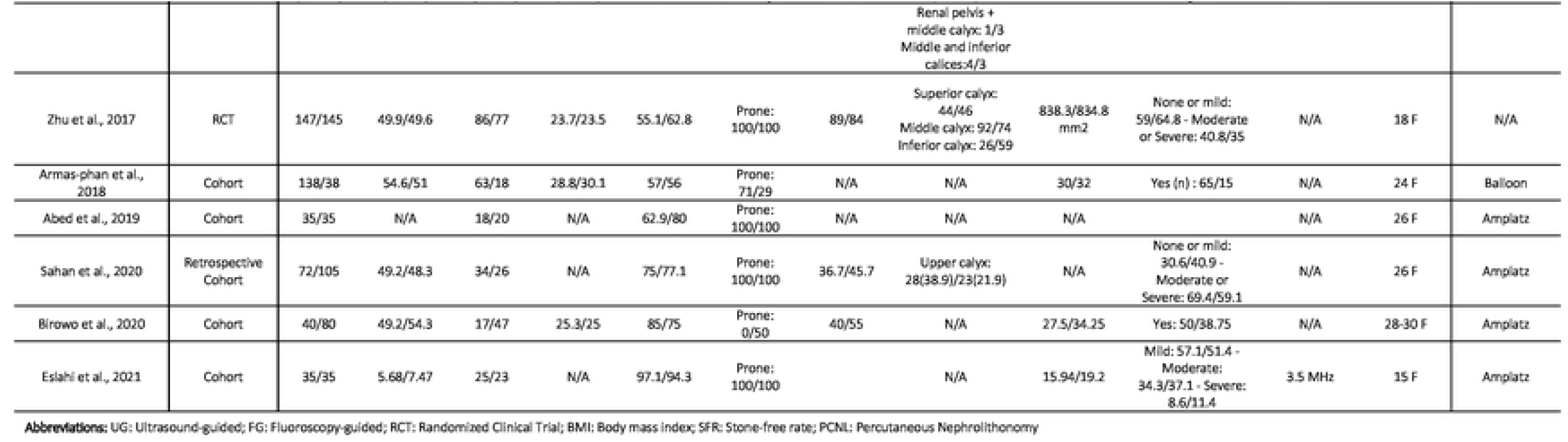
Characteristics of Included Studies

### Primary outcomes SFR

Eighteen studies comprising 2815 patients (of which 1411 underwent UG-PCNL) were included in our analysis (12-29). No significant differences were observed in the case of SFR between ultrasound and fluoroscopy-guided patients (RR: 1.02; 95% CI: 0.98 to 1.06; p = 0.29; I2 = 46%) (Fig. 2-A). Furthermore, no significant between-group differences were observed based on the study design (p = 0.75). Our meta-regression analysis revealed no significant association between publication year (as a measure of clinicians’ experience) and risk of SFR (p= 0.81) (Fig. 5). Visual inspection of the funnel plot revealed no possible source of small study effects (Fig. 3-A). This was further confirmed by using the Eggers regression test for funnel plot asymmetry (p= 0.40).

**Fig. 2:**
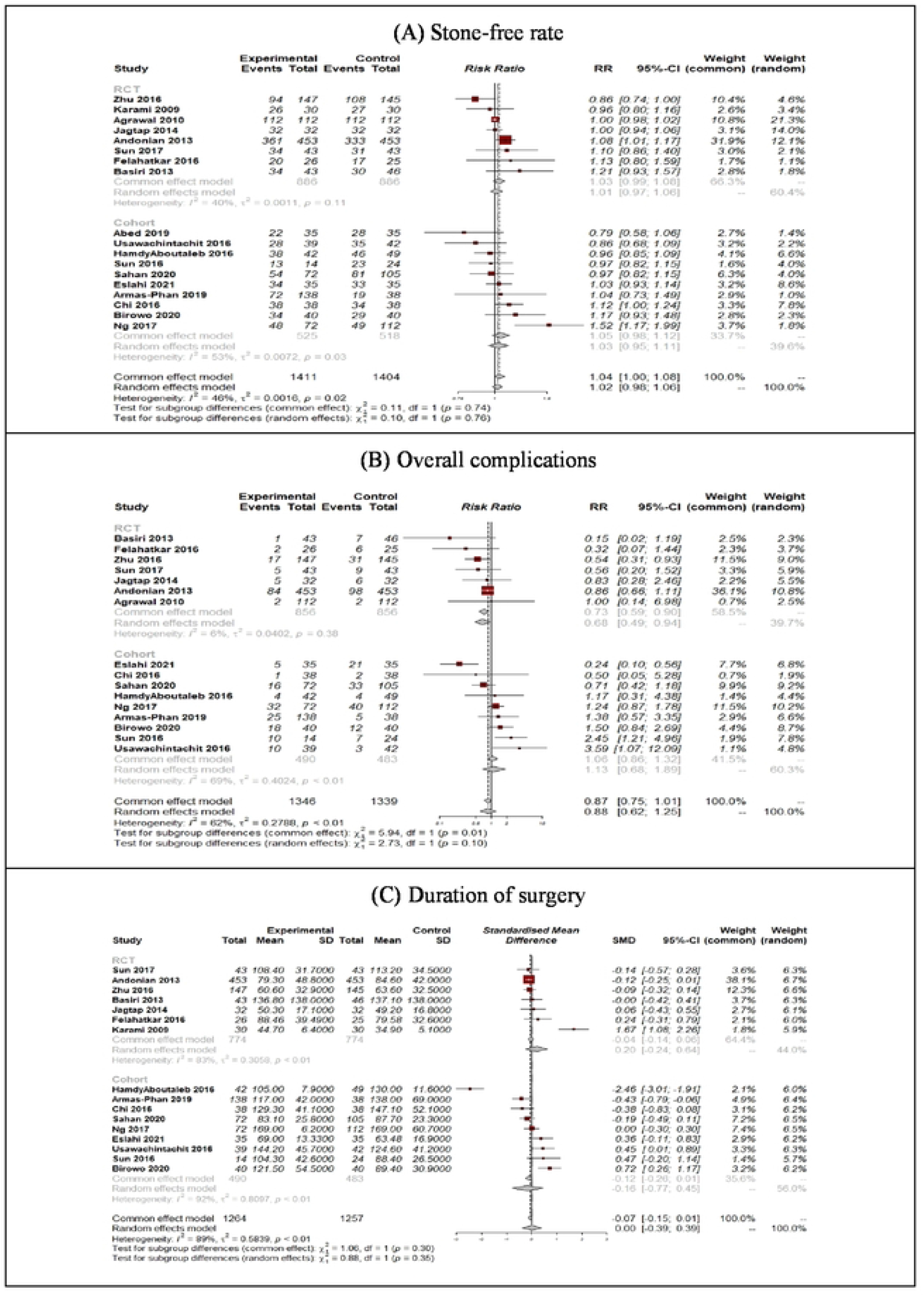
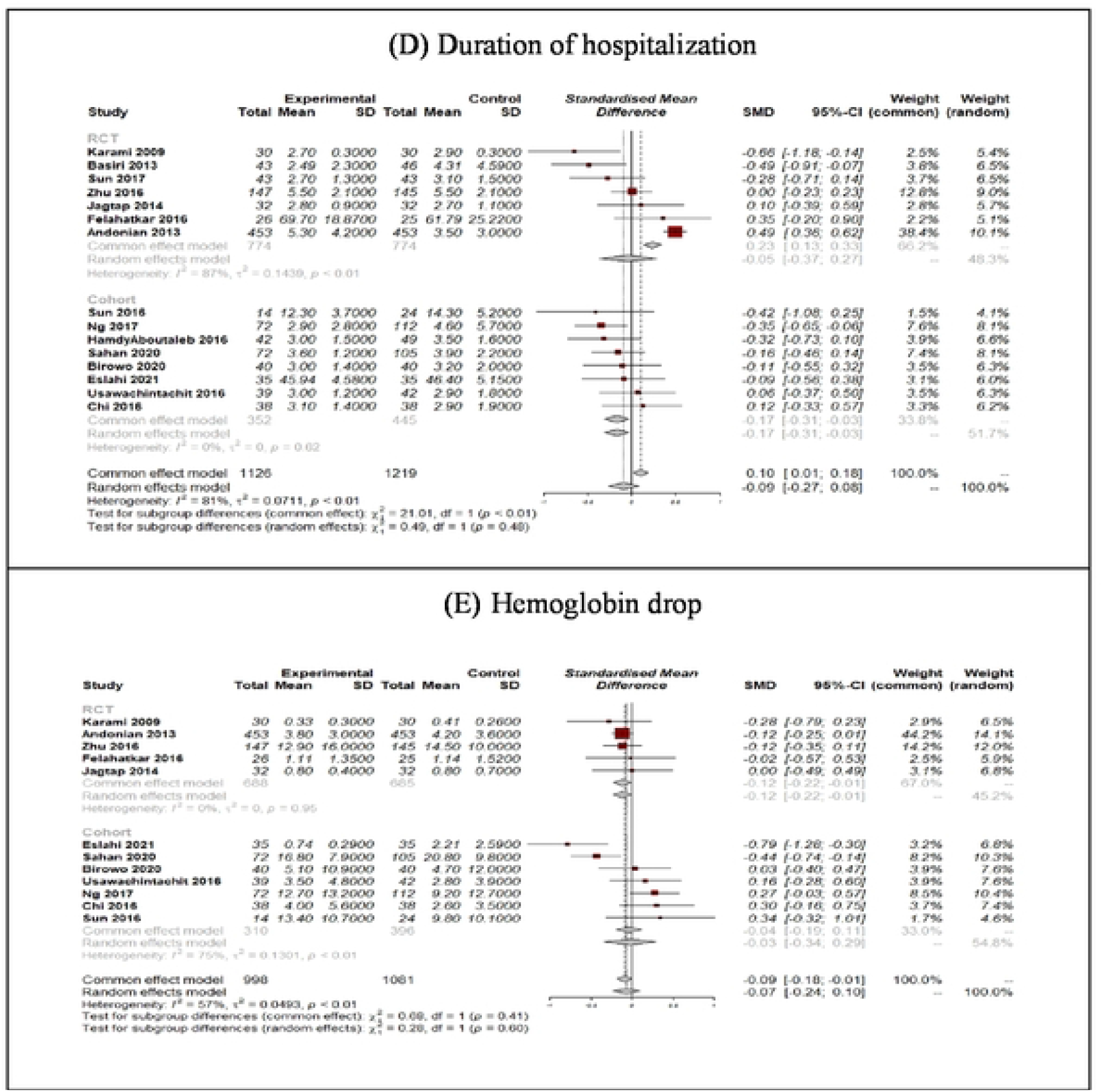
Forest plots of primary outcomes of UG-PCNL (experimental) versus FG-PCNL (control)

**Fig. 3.**
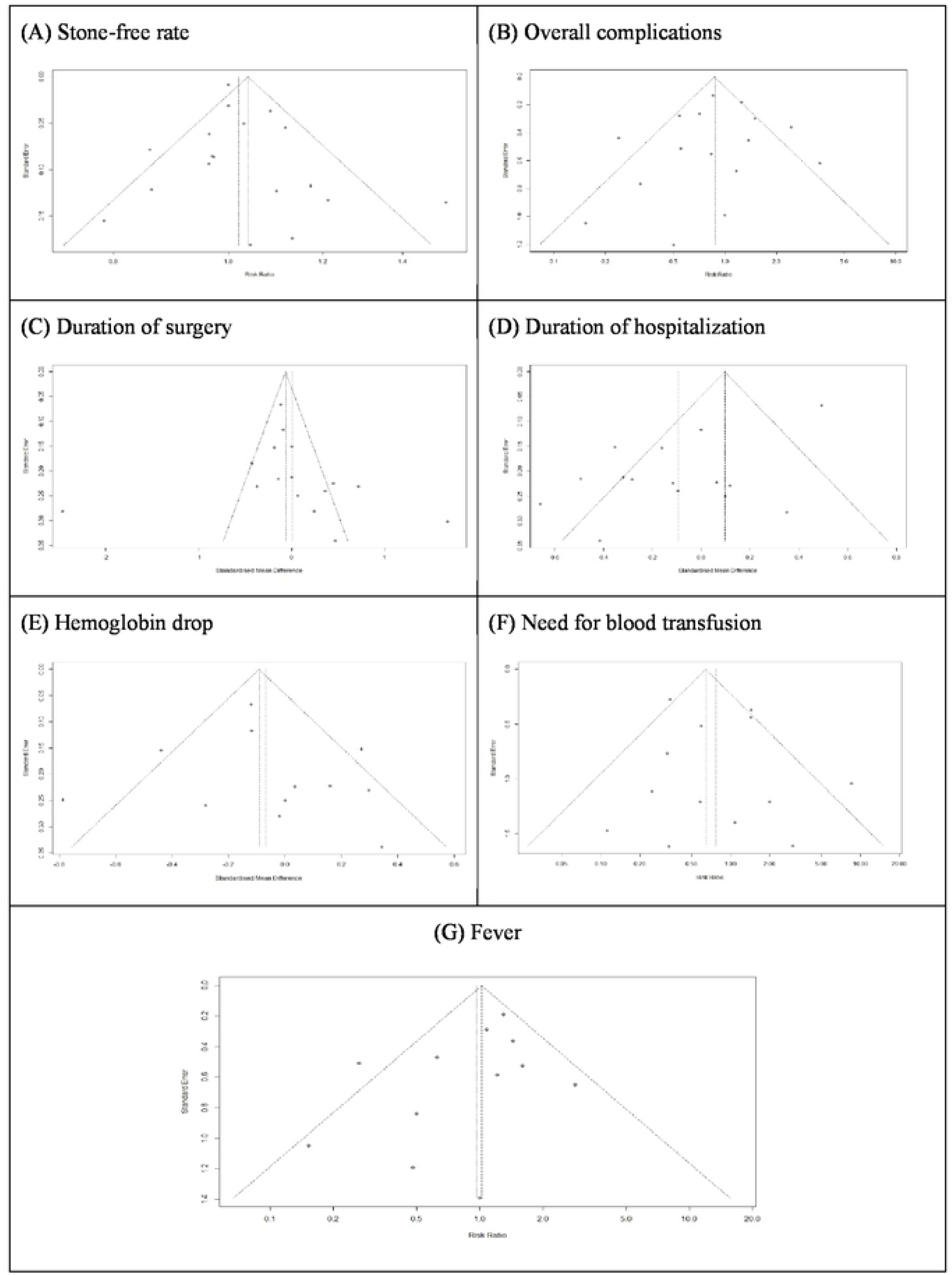
Funnel plots of outcomes reported in at least 10 studies

**Fig. 4.**
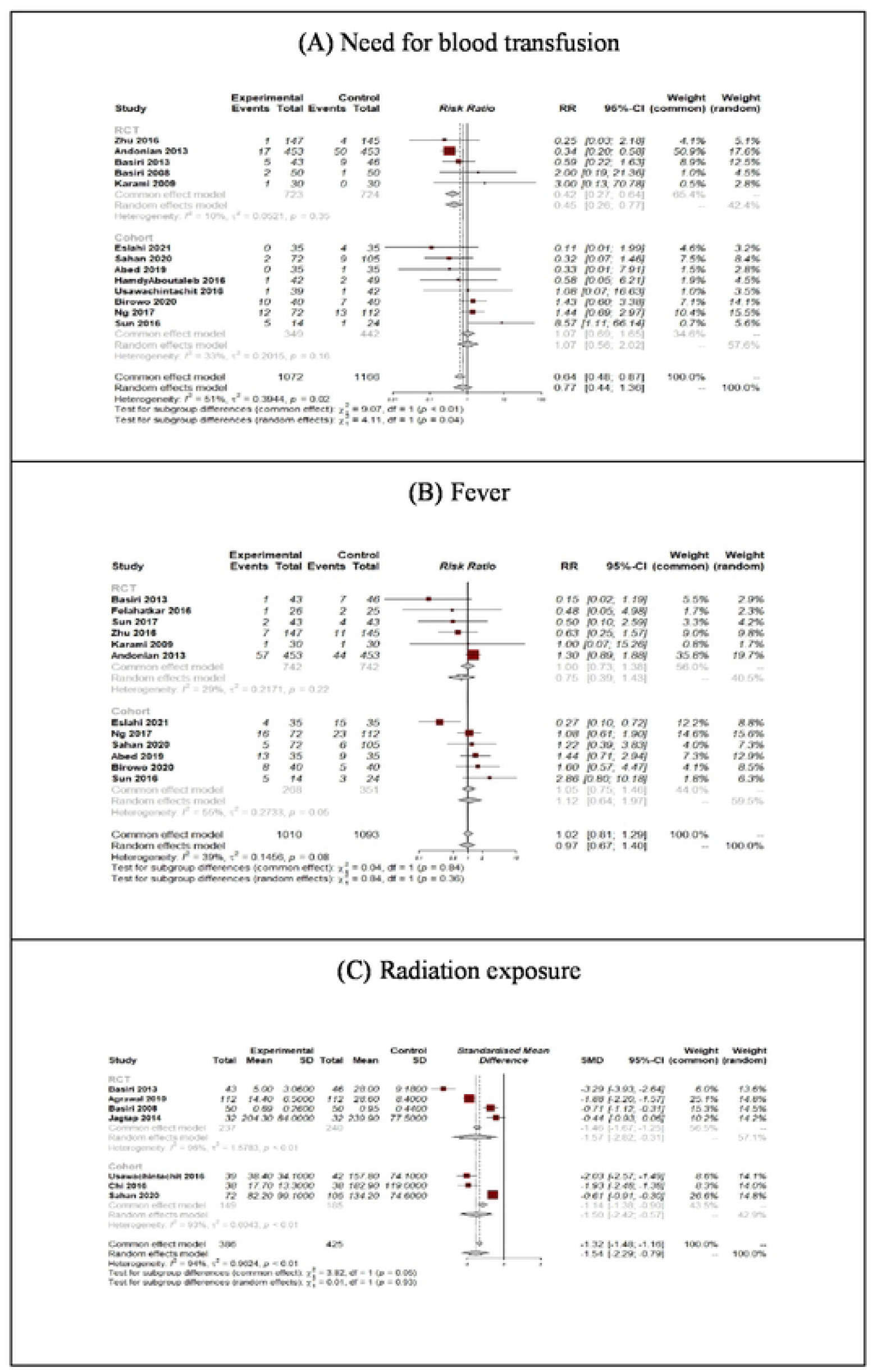
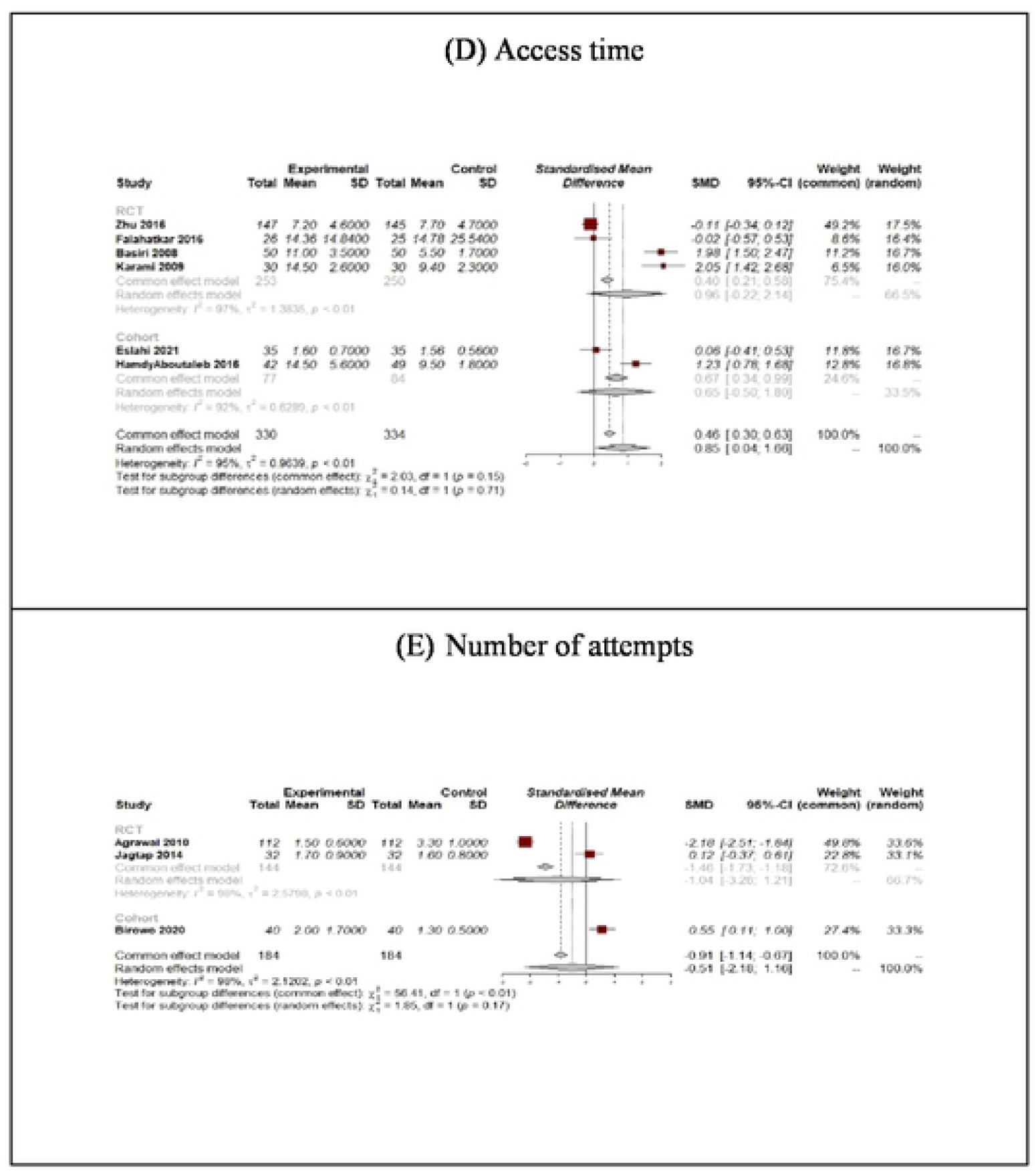
Forest plots of secondary outcomes ofUG-PCNL (experimental) versus FG-PCNL (control)

**Fig. 5.**
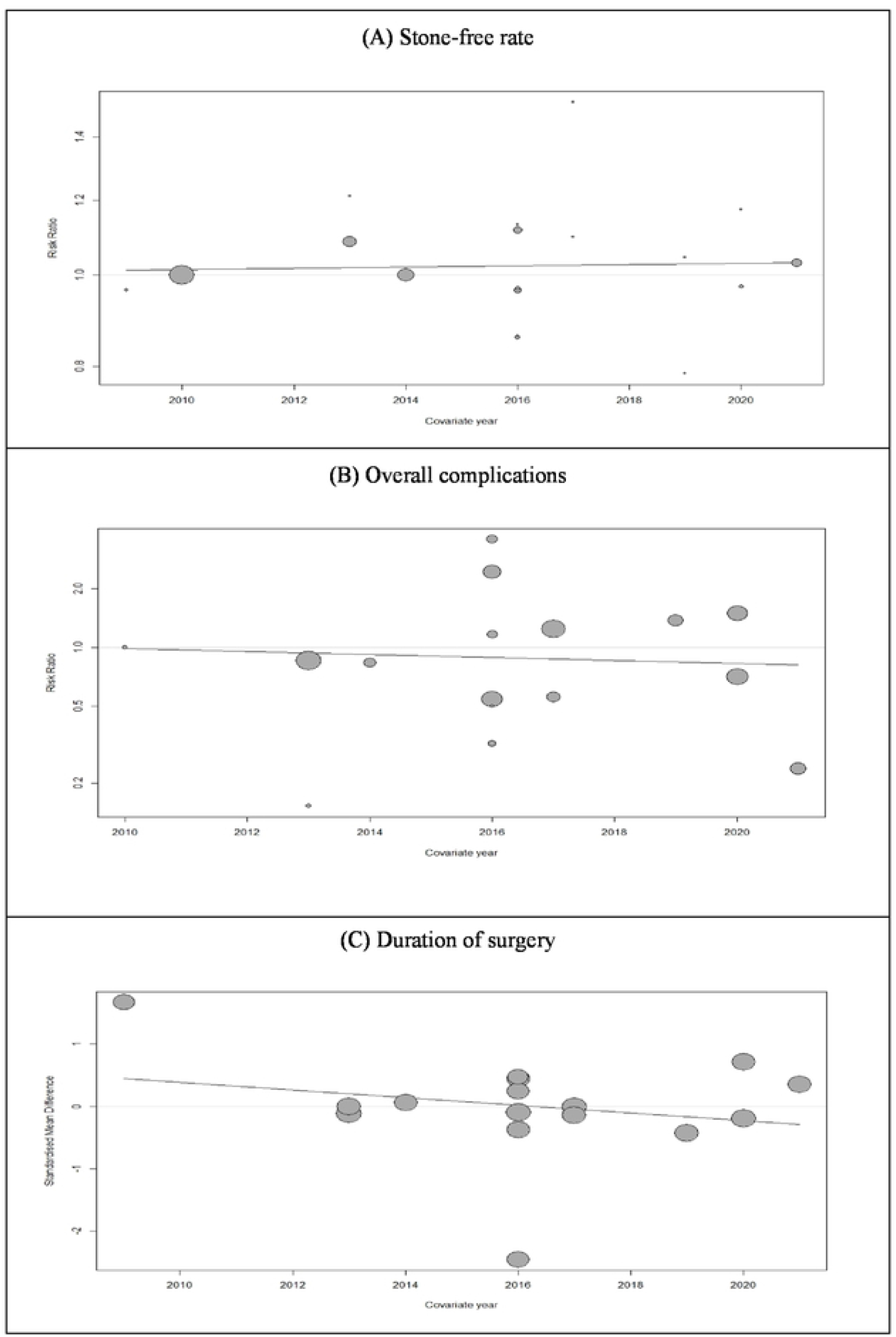
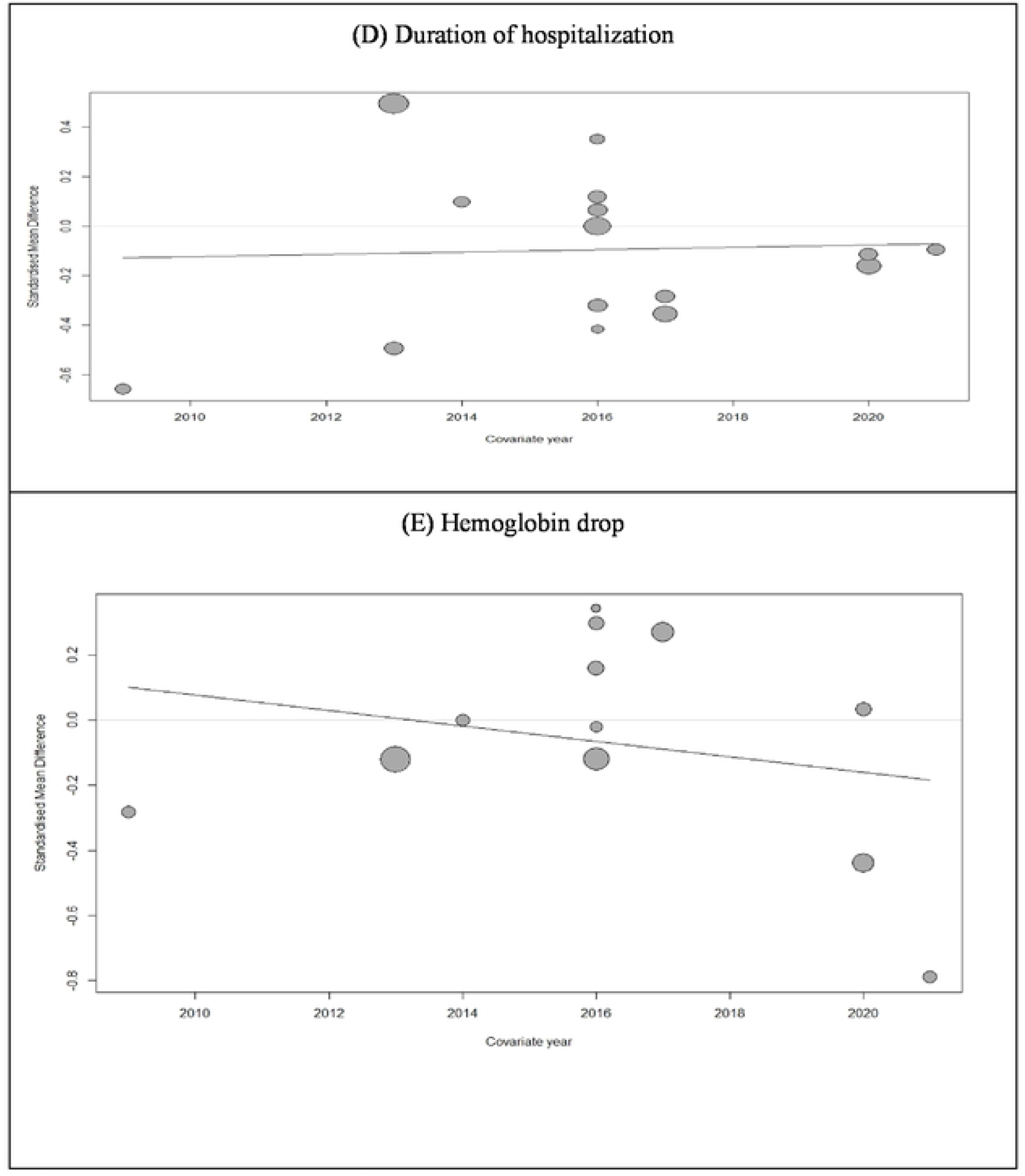
Results of meta-regression. Effects of publication year and the experience of the clinician on primary outcomes

### Overall complication

For comparing the rate of complications between groups, studies reporting the outcome of the Clavien-Dindo classification system were included in this analysis. Sixteen studies reported compared complications based on this classification (13-24, 26-29). These studies included 1346 and 1339 patients treated with ultrasound and fluoroscopy-guided PCNL, respectively. Meta-analysis of these studies revealed no significant difference in overall complication (RR: 0.88; 95% CI: 0.62 to 1.25; p = 0.47; I2 = 62%) (Fig. 2-B). However, the results of our subgroup analysis showed a significant reduction in the overall complication rate in the ultrasound-guided group when pooling the results of RCTs independently (RR: 0.73; 95% CI: 0.59 to 0.90; I2 = 6%). No significant association between publication year and complication was observed (p= 0.80) (Fig. 5). Visual inspection of the funnel plot revealed no possible source of small study effects (Fig. 3-B). This was further confirmed by using the Eggers regression test for funnel plot asymmetry (p= 0.62).

### Duration of surgery

Sixteen studies, including 2521 patients (of which 1264 underwent ultrasound-guided PCNL), were included in our analysis (12, 14-24, 26-29). No significant differences were observed in operation time between ultrasound and fluoroscopy-guided patients (SMD: 0.00; 95% CI: -0.39 to 0.39; p = 0.98; I2 = 89%) (Fig. 2-C). No significant between-group differences were observed based on the study design (p = 0.35). Our meta-regression revealed a reduced SMD by increasing publication year (as a measure of clinicians’ experience) (Fig. 5). Although this interpretation was made based on a non-significant result (p= 0.37). Visual inspection of the funnel plot revealed no possible source of small study effects (Fig. 3-C). This was further confirmed by using the Eggers regression test for funnel plot asymmetry (p= 0.58).

### Duration of hospitalization

Sixteen studies comprising 2345 patients (of which 1126 underwent ultrasound-guided PCNL) were included in our analysis (12, 14-24, 27-29). No significant differences were observed in duration of hospitalization between ultrasound and fluoroscopy-guided patients (SMD: -0.09; 95% CI: -0.27 to 0.08; p = 0.28; I2 = 81%) (Fig. 2-D). The pooled effect size from observational studies showed a favorable outcome in ultrasound-guided patients (SMD: -0.17; 95% CI: -0.31 to -0.03; I2 = 0%). Our meta-regression revealed a non-significant association between publication year and duration of hospitalization (p= 0.88) (Fig. 5). Visual inspection of the funnel plot revealed the presence of small study effects as an indication of publication bias (Fig. 3-D). This was further confirmed by using the Eggers regression test for funnel plot asymmetry (p= 0.001).

### Hb drop

Twelve studies comprising 2079 patients (of which 998 underwent ultrasound-guided PCNL) were included (12, 14, 16-18, 20-22, 24, 27-29). Overall, no significant differences were observed in Hb drop between ultrasound and fluoroscopy-guided patients after their operation (SMD: -0.07; 95% CI: -0.24 to 0.10; p = 0.42; I2 = 57%) (Fig. 2-E). The pooled results of RCTs showed a favorable outcome in ultrasound-guided patients (SMD: -0.12; 95% CI: -0.22 to -0.01; I2 = 0%). Our meta-regression revealed a non-significant association between publication year and duration of hospitalization (p= 0.41) (Fig. 5). Visual inspection of the funnel plot revealed no possible source of small study effects (Fig. 3-E). This was further confirmed by using the Eggers regression test for funnel plot asymmetry (p= 0.65).

### Secondary outcomes

#### Need for blood transfusion

Thirteen studies with 2238 patients (of which 998 underwent UG-PCNL) were included (12, 14, 15, 19-22, 24, 25, 27-30). Overall, no significant differences were observed in blood transfusion rate between ultrasound and fluoroscopy-guided patients (RR: 0.77; 95% CI: 0.44 to 1.36; p = 0.37; I2 = 51%) (Fig. 4-A). The pooled results of RCTs showed a reduced need for blood transfusion in ultrasound-guided patients (RR: 0.42; 95% CI: 0.27 to 0.64; I2 = 10%). Visual inspection of the funnel plot revealed no possible source of small study effects (Fig. 3-F). This was further confirmed by using the Eggers regression test for funnel plot asymmetry (p= 0.55).

#### Fever

Twelve studies with 2103 patients (of which 1010 underwent UG-PCNL) were included (12, 14, 15, 18, 21-25, 27-29). No significant differences were observed between ultrasound and fluoroscopy-guided patients (RR: 0.97; 95% CI: 0.67 to 1.40; p = 0.87; I2 = 39%) (Fig. 4-B). Our subgroup analysis showed no significant differences between the results of RCTs and observational studies (Between groups p= 0.36). Visual inspection of the funnel plot revealed no possible source of small study effects (Fig. 3-G). This was further confirmed by using the Eggers regression test for funnel plot asymmetry (p= 0.17).

#### Radiation exposure

Seven studies comprising 811 patients (of which 386 underwent UG-PCNL) were included (13, 15-17, 20, 28, 30). When comparing the duration for which patients were exposed to radiation, significant differences between ultrasound-guided patients and fluoroscopy-guided patients were found. (SMD: -1.54; 95% CI: -2.29 to -0.79; p < 0.0001; I2 = 94%) (Fig. 4-C).

#### Access time

Six studies, including 664 patients (of which 330 underwent UG-PCNL), were included (12, 18, 19, 24, 29, 30). Our analysis revealed a shorter time needed for having access through the fluoroscopy-guided technique. (SMD: 0.85; 95% CI: 0.04 to 1.66; p= 0.04; I2 = 95%) (Fig. 4-D).

#### Number of attempts

Three studies, including 368 patients (of which 184 underwent UG-PCNL), were included (13, 16, 27). No significant differences were observed between ultrasound and fluoroscopy-guided patients (SMD: -0.51; 95% CI: -2.18 to 1.16; p= 0.54; I2 = 98%) (Fig. 4-E).

## Discussion

To remove the renal stones, PCNL is offered with the highest SFR alongside high complication(31). SFR is defined as the absence of residual stones, or the presence of residual stone fragments less than 4 mm in size in follow-up studies such as kidney ultrasound and non-contrast computed tomography (CT) (32).

The indications for PCNL are stones larger than 20 mm, staghorn, and partial staghorn calculi(33). The contraindications for PCNL include pregnancy, bleeding disorders, and uncontrolled urinary tract infections(34).

PCNL is an efficient technique for removing large and complex stones with high success and low morbidity rates(35). PCNL is performed by making a small incision in the flank area under fluoroscopy or ultrasound guidance(36). PCNL under ultrasound guidance has some advantages, including the absence of ionizing radiation, shorter time of the procedure, less puncture, and no more use of contrast agents(37-39).To decrease complications like blood loss, postoperative pain, and renal damage due to larger instruments, a modification of the standard procedure was propounded (41). In fluoroscopy-guided PCNL, contrast is injected through a urethral catheter. Puncture failure and re-do FG-PCNL can be mentioned as a disadvantage of this procedure(40, 41). Some studies mention that UG-PCNL in the flank or the prone position has a higher success rate and fewer complications compared with FG-PCNL(7, 42). In addition, mini-PCNL is a standard technique in the treatment of renal and upper ureteric stones using a 28–30 F ureteroscope(43).

In 2018, Yu-Hsiang and his colleagues published a meta-analysis comparing UG-PCNL with FG-PCNL(44). It had eight included articles, and based on their analysis, they reported that UG-PCNL had a significantly lower complication rate and also fewer intraoperative complications than FG-PCNL(45, 46). Accordingly, UG-PCNL was also associated with reduced inadvertent organ injury risks. Therefore, it can be concluded that ultrasound provides information about the surrounding viscera, determines the depth of needle penetration, and identifies the area posterior to the anterior calyces when comparing the different complication rates(47). Due to its analysis, UG-PCNL in the supine position had a higher SFR and significantly lower complication rate than FG-PCNL in the supine position. According to its results, it did not imply statistically significant differences in SFR. however, it mentioned if a patient is an appropriate case for a supine position PCNL, like patients with cardiovascular disease or spinal deformities, UG-PCNL can be a better choice than FG-PCNL(11, 48). In mini-PNCL, they perform with a smaller percutaneous tract by using a miniature endoscope. Accordingly, mini-PCNL was associated with less bleeding and postoperative pain during the procedure(49, 50).

We conducted a systematic review and meta-analysis of efficacy and safety in UG-PCNL versus FG-PCNL with 19 articles included. This study was the first meta-analysis to evaluate ultrasound versus fluoroscopy as imaging guidance for percutaneous nephrolithotomy, including RCTs and observational Cohort studies. The sample size of the articles ranged from 45 to 906. A total of 3016 patients were evaluated in the included studies (of which 1521 patients underwent UG-PCNL). A regression analysis was done based on publication year and approaching recent years, with increasing experience in performing UG-PCNL. A better result was seen than FG-PCNL. Outcomes were considered in this meta-analysis in two groups. Each group had five sub-groups. Primary outcomes included SFR, overall complications, duration of surgery, duration of hospitalization, and Hb drop. Secondary outcomes considered the need for blood transfusion, fever, radiation exposure, access time, and the number of attempts. In most outcomes except radiation exposure and access time, no significant differences were observed between ultrasound and fluoroscopy-guided studies. Based on our analysis, RCT studies presented fewer complication rates compared to Cohort studies. In summary, comparing the results of this study to the systematic review and meta-analysis published in 2018, most outcomes did not display significant differences. Generally, between these two methods considered in this study, the results were in favor of UG-PCNL.

### Limitations

There are multiple reasons for the heterogeneity of the included studies and also the conflicting results. On the one hand, the patients had different baseline characteristics, including body mass index and hydronephrosis degree. On the other hand, the procedure was conducted in various positions. In addition, ultrasound is operator-dependent, and its practicality is based on machine properties and operator expertise.

### Conclusion

According to the less radiation exposure when using the UG-PCNL technique, we suggest prioritizing the utilization of UG-PCNL in treating renal stones. Regarding the fact that the experience and level of expertise of the ultrasound performer may affect the findings, it is recommended to prepare a program to train people who perform the procedure. This could reduce the errors, which are mainly due to the lack of organized training programs and sufficient skills.

## Data Availability

The data underlying the results presented in the study are available from the email address of the corresponding author

## Acknowledgments

We thank all the authors of the included studies and all those who contributed to the present study.

## Author Contributions

**Conceptualization:** Razman Arabzadeh Bahri.

**Data curation:** Razman Arabzadeh Bahri, Saba Maleki, Arman Shafiee.

**Formal analysis:** Arman Shafiee.

**Investigation:** Razman Arabzadeh Bahri, Saba Maleki.

**Methodology:** Arman Shafiee.

**Supervision:** Parnian Shobeiri.

**Visualization:** Razman Arabzadeh Bahri.

**Writing – original draft:** Razman Arabzadeh Bahri, Saba Maleki, Arman Shafiee.

**Writing – review & editing:** Razman Arabzadeh Bahri, Saba Maleki, Arman Shafiee, Parnian Shobeiri.

## References

1. Ziemba JB, Matlaga BR. Epidemiology and economics of nephrolithiasis. Investigative and clinical urology. 2017;58(5):299–306.

2. Türk C, Petřík A, Sarica K, Seitz C, Skolarikos A, Straub M, et al. EAU guidelines on interventional treatment for urolithiasis. European urology. 2016;69(3):475–82.

3. Assimos D, Krambeck A, Miller NL, Monga M, Murad MH, Nelson CP, et al. Surgical management of stones: American urological association/endourological society guideline, PART I. The Journal of urology. 2016;196(4):1153–60.

4. Hidajat N, Wust P, Kreuschner M, Felix R, Schroder R. Radiation risks for the radiologist performing transjugular intrahepatic portosystemic shunt (TIPS). The British Journal of Radiology. 2006;79(942):483–6.

5. Vano E, Kleiman NJ, Duran A, Romano-Miller M, Rehani MM. Radiation-associated lens opacities in catheterization personnel: results of a survey and direct assessments. Journal of Vascular and Interventional Radiology. 2013;24(2):197–204.

6. Mancini JG, Raymundo EM, Lipkin M, Zilberman D, Yong D, Bañez LL, et al. Factors affecting patient radiation exposure during percutaneous nephrolithotomy. The Journal of urology. 2010;184(6):2373–7.

7. Hosseini MM, Hassanpour A, Farzan R, Yousefi A, Afrasiabi MA. Ultrasonography-guided percutaneous nephrolithotomy. Journal of endourology. 2009;23(4):603–7.

8. Hudnall M, Usawachintachit M, Metzler I, Tzou DT, Harrison B, Lobo E, et al. Ultrasound guidance reduces percutaneous nephrolithotomy cost compared to fluoroscopy. Urology. 2017;103:52–8.

9. Ahmad AA, Alhunaidi O, Aziz M, Omar M, Al-Kandari AM, El-Nahas A, et al. Current trends in percutaneous nephrolithotomy: an internet-based survey. Therapeutic Advances in Urology. 2017;9(9-10):219–26.

10. Kwee MM, Ho Y-H, Rozen WM. The prone position during surgery and its complications: a systematic review and evidence-based guidelines. International surgery. 2015;100(2):292–303.

11. Karaolides T, Moraitis K, Bach C, Masood J, Buchholz N. Positions for percutaneous nephrolithotomy: Thirty-five years of evolution. Arab Journal of Urology. 2012;10(3):307–16.

12. Karami H, Rezaei A, Mohammadhosseini M, Javanmard B, Mazloomfard M, Lotfi B. Ultrasonography-guided percutaneous nephrolithotomy in the flank position versus fluoroscopy-guided percutaneous nephrolithotomy in the prone position: a comparative study. J Endourol. 2010;24(8):1357–61.

13. Agarwal M, Agrawal MS, Jaiswal A, Kumar D, Yadav H, Lavania P. Safety and efficacy of ultrasonography as an adjunct to fluoroscopy for renal access in percutaneous nephrolithotomy (PCNL). BJU Int. 2011;108(8):1346–9.

14. Andonian S, Scoffone CM, Louie MK, Gross AJ, Grabe M, Daels FP, et al. Does imaging modality used for percutaneous renal access make a difference? A matched case analysis. J Endourol. 2013;27(1):24–8.

15. Basiri A, Mirjalili MA, Kardoust Parizi M, Moosa Nejad NA. Supplementary X-ray for ultrasound-guided percutaneous nephrolithotomy in supine position versus standard technique: a randomized controlled trial. Urol Int. 2013;90(4):399–404.

16. Jagtap J, Mishra S, Bhattu A, Ganpule A, Sabnis R, Desai MR. Which is the preferred modality of renal access for a trainee urologist: ultrasonography or fluoroscopy? Results of a prospective randomized trial. J Endourol. 2014;28(12):1464–9.

17. Chi T, Masic S, Li J, Usawachintachit M. Ultrasound Guidance for Renal Tract Access and Dilation Reduces Radiation Exposure during Percutaneous Nephrolithotomy. Adv Urol. 2016;2016:3840697.

18. Falahatkar S, Allahkhah A, Kazemzadeh M, Enshaei A, Shakiba M, Moghaddas F. Complete supine PCNL: ultrasound vs. fluoroscopic guided: a randomized clinical trial. Int Braz J Urol. 2016;42(4):710–6.

19. HamdyAboutaleb M, Mohammed El-Shazly MJAS. Ultrasound versus fluoroscopy guided percutaneous nephrolithotomy for treatment of calculi in hydronephrotic kidneys. 2016;2:015.

20. Usawachintachit M, Masic S, Chang HC, Allen IE, Chi T. Ultrasound Guidance to Assist Percutaneous Nephrolithotomy Reduces Radiation Exposure in Obese Patients. Urology. 2016;98:32–8.

21. Ng FC, Yam WL, Lim TYB, Teo JK, Ng KK, Lim SK. Ultrasound-guided percutaneous nephrolithotomy: Advantages and limitations. Investig Clin Urol. 2017;58(5):346–52.

22. Sun H, Zhang Z, Huang G, Wan SP, Chen H, He B, et al. Fluoroscopy versus ultrasonography guided mini-percutaneous nephrolithotomy in patients with autosomal dominant polycystic kidney disease. Urolithiasis. 2017;45(3):297–303.

23. Sun W, Liu MN, Yang ZW, Wang Q, Xu Y. Ultrasound-guided percutaneous nephrolithotomy for the treatment in patients with kidney stones. Medicine (Baltimore). 2017;96(51):e9232.

24. Zhu W, Li J, Yuan J, Liu Y, Wan SP, Liu G, et al. A prospective and randomised trial comparing fluoroscopic, total ultrasonographic, and combined guidance for renal access in mini-percutaneous nephrolithotomy. BJU Int. 2017;119(4):612–8.

25. Abed SM, Alhamdani NJIJMS, 18. Ultrasonographic guidance versus fluoroscopic guidance for renal access in percutaneous nephrolithotomy (PCNL): a comparative study. 2019;335.

26. Armas-Phan M, Tzou DT, Bayne DB, Wiener SV, Stoller ML, Chi T. Ultrasound guidance can be used safely for renal tract dilatation during percutaneous nephrolithotomy. BJU Int. 2020;125(2):284–91.

27. Birowo P, Raharja PAR, Putra HWK, Rustandi R, Atmoko W, Rasyid N. X-ray-free ultrasound-guided versus fluoroscopy-guided percutaneous nephrolithotomy: a comparative study with historical control. Int Urol Nephrol. 2020;52(12):2253–9.

28. Sahan A, Cubuk A, Ozkaptan O, Ertas K, Canakci C, Eryildirim B, et al. Safety of Upper Pole Puncture in Percutaneous Nephrolithotomy with the Guidance of Ultrasonography versus Fluoroscopy: A Comparative Study. Urol Int. 2020;104(9-10):769–74.

29. Eslahi A, Ahmed F, Hosseini MM, Rezaeimehr MR, Fathi N, Nikbakht HA, et al. Minimal invasive percutaneous nephrolithotomy (Mini-PCNL) in children: Ultrasound versus fluoroscopic guidance. Arch Ital Urol Androl. 2021;93(2):173–7.

30. Basiri A, Ziaee AM, Kianian HR, Mehrabi S, Karami H, Moghaddam SM. Ultrasonographic versus fluoroscopic access for percutaneous nephrolithotomy: a randomized clinical trial. J Endourol. 2008;22(2):281–4.

31. Assimos D, Krambeck A, Miller NL, Monga M, Murad MH, Nelson CP, et al. Surgical management of stones: American urological association/endourological society guideline, PART I. 2016;196(4):1153–60.

32. Atis G, Culpan M, Pelit ES, Canakci C, Ulus I, Gunaydin B, et al. Comparison of Percutaneous Nephrolithotomy and Retrograde Intrarenal Surgery in Treating 20-40 mm Renal Stones. Urol J. 2017;14(2):2995–9.

33. Desai M, Sun Y, Buchholz N, Fuller A, Matsuda T, Matlaga B, et al. Treatment selection for urolithiasis: percutaneous nephrolithomy, ureteroscopy, shock wave lithotripsy, and active monitoring. World journal of urology. 2017;35(9):1395–9.

34. Preminger GM, Assimos DG, Lingeman JE, Nakada SY, Pearle MS, Wolf JS. Chapter 1: AUA guideline on management of staghorn calculi: diagnosis and treatment recommendations. The Journal of urology. 2005;173(6):1991–2000.

35. Sun W, Liu M-n, Yang Z-w, Wang Q, Xu Y. Ultrasound-guided percutaneous nephrolithotomy for the treatment in patients with kidney stones. Medicine. 2017;96(51).

36. Fernström I, Johansson B. Percutaneous pyelolithotomy: a new extraction technique. Scandinavian journal of urology and nephrology. 1976;10(3):257–9.

37. Zegel H, Pollack H, Banner M, Goldberg B, Arger P, Mulhern C, et al. Percutaneous nephrostomy: comparison of sonographic and fluoroscopic guidance. American Journal of Roentgenology. 1981;137(5):925–7.

38. Basiri A, Ziaee SAM, Nasseh H, Kamranmanesh M, Masoudy P, Heidary F, et al. Totally ultrasonography-guided percutaneous nephrolithotomy in the flank position. Journal of endourology. 2008;22(7):1453–8.

39. Gupta S, Gulati M, Suri S. Ultrasound-guided percutaneous nephrostomy in non-dilated pelvicaliceal system. Journal of clinical ultrasound. 1998;26(3):177–9.

40. HamdyAboutaleb M, Mohammed El-Shazly M. Ultrasound versus fluoroscopy guided percutaneous nephrolithotomy for treatment of calculi in hydronephrotic kidneys. Ann SurgInt. 2016;2:015.

41. Inanloo SH, Yahyazadeh SR, Rashidi S, Amini E, Nowroozi MR, Ayayti M, et al. Feasibility and safety of ultrasonography guidance and flank position during percutaneous nephrolithotomy. 2018;200(1):195–201.

42. Basiri A, Ziaee AM, Kianian HR, Mehrabi S, Karami H, Moghaddam SMH. Ultrasonographic versus fluoroscopic access for percutaneous nephrolithotomy: a randomized clinical trial. Journal of endourology. 2008;22(2):281–4.

43. Desai J, Zeng G, Zhao Z, Zhong W, Chen W, Wu W. A novel technique of ultra-mini-percutaneous nephrolithotomy: introduction and an initial experience for treatment of upper urinary calculi less than 2 cm. BioMed research international. 2013;2013.

44. Yang Y-H, Wen Y-C, Chen K-C, Chen C. Ultrasound-guided versus fluoroscopy-guided percutaneous nephrolithotomy: a systematic review and meta-analysis. World journal of urology. 2019;37(5):777–88.

45. Ng FC, Yam WL, Lim TYB, Teo JK, Ng KK, Lim SK. Ultrasound-guided percutaneous nephrolithotomy: advantages and limitations. Investigative and clinical urology. 2017;58(5):346–52.

46. Wang K, Zhang P, Xu X, Fan M. Ultrasonographic versus fluoroscopic access for percutaneous nephrolithotomy: a meta-analysis. Urologia internationalis. 2015;95(1):15–25.

47. Lojanapiwat B. The ideal puncture approach for PCNL: Fluoroscopy, ultrasound or endoscopy? Indian journal of urology: IJU: journal of the Urological Society of India. 2013;29(3):208.

48. Manohar T, Jain P, Desai M. Supine percutaneous nephrolithotomy: effective approach to high-risk and morbidly obese patients. Journal of endourology. 2007;21(1):44–9.

49. Jackman SV, Docimo SG, Cadeddu JA, Bishoff JT, Kavoussi LR, Jarrett TW. The “mini-perc” technique: a less invasive alternative to percutaneous nephrolithotomy. World journal of urology. 1998;16(6):371–4.

50. Haghighi R, Zeraati H, Zade MG. Ultra-mini-percutaneous nephrolithotomy (PCNL) versus standard PCNL: A randomised clinical trial. Arab Journal of Urology. 2017;15(4):294–8.

